# Challenges and Health Disparities in Prostate Cancer: Healthcare Access as a Driver of Inequities in a Rural Hospital Network

**DOI:** 10.64898/2026.11.05.26367368

**Authors:** Antoni Aguilar-Mogas, J. Nathaniel Hamilton, Andrew W. Ju, C. Suzanne Lea, Charles H. Shelton

## Abstract

**Background:** Prostate cancer is the most common male cancer, and second most common cause of cancer mortality in the US. Disparities abound in this disease for reasons that need further study, including social and ethnic barriers, geographic barriers, and biological differences. North Carolina is the second largest state by rural population measures (3.5 million), which provides ample geography to study these disparities.

**Materials and Methods:** A dataset containing all patients with a medical record documenting a diagnosis of and/or treatment for de novo prostate cancer within the mostly rural ECU Health 9-hospital system was characterized over a 20-year period for structural inequities, access barriers, and biological differences.

**Results:** Stages and outcomes of prostate cancer changed temporally, reflecting changes in screening, treatment, and inclusion of prognostic factors such as prostate-specific antigen, Gleason score, and other factors. Late stage (4) prostate cancer disproportionately affected our system as a whole and smaller community hospitals, with higher death rates seen in community, mostly rural programs that lack access to all required components of care.

**Conclusion:** Stages of prostate cancer changed considerably over the study period, reflecting changes in screening recommendations, and staging criteria. Late stage (4) disease increased over the analytic period, was higher than NCDB references by period, and disproportionately affected community programs where resources were sparse, correlating with worse outcomes.

## INTRODUCTION

Prostate cancer (PCa) is the most common cancer and the second leading cause of cancer-related deaths among men in the US ^1^. While PCa mortality rates have declined over the last few decades, these improvements are not proportional across populations groups ^2^. PCa has the largest racial disparity of any cancer, with Black men diagnosed at a higher rate than White men in the US, and twice as likely to die from it ^3^. These differences likely relate to structural inequities, uneven access to quality care, genetic/biological differences, and other factors ^4^.

Mortality from PCa is preventable ^5^ given the existence of screening and early detection techniques ^6^ and its relation to lifestyle and environmental factors ^7^ . The most common approach to identify at-risk patients for PCa detection is prostate-specific antigen (PSA) testing ^8^. However, screening for PCa using PSA alone is controversial due to a potential risk of over diagnosis and overtreatment of indolent disease ^9^. Current American Urological Association guidelines recommend using PSA as the first screening test, offering the screening to all men aged 50 to 69 years, and starting at 40 or 45 years for men at increased risk of PCa. Increased risk men include: Black men, men with known mutations, or men with family history of PCa ^6^.

Risk factors including environmental conditions, personal actions or habits, and family history increase an individual’s likelihood of developing cancer ^10^. In addition, individuals living in economically distressed areas experience higher cancer mortality rates, partly due to reduced access to providers, diagnostics, and treatments ^11^. North Carolina (NC) is the ninth largest state by total population, and second largest by rural population volume ^12^. Eastern North Carolina (ENC), the easternmost region of the state, represents a mostly rural and socially vulnerable region: poverty levels in ENC are higher than both state and national averages, where median income and educational attainment are low ^13^. Consequently, health indicators in ENC reflect significant health disparities: from life expectancy ^14^ to cancer incidence and mortality ^15^ and overall experience of healthcare ^16^.

ECU Health is the largest healthcare system in ENC, providing service to 29 counties through a hub-in-spoke model of care involving nine hospitals: ECU Health Medical Center serves as a central urban hub, with eight community hospitals serving as rural providers of some urological cancer care. Because few programs contain all needed providers, patients face additional barriers to access care including logistical challenges of transportation, lodging, support and navigation. This study reviews clinical outcomes for all patients diagnosed or treated in a single health care network to understand factors that link outcomes to social vulnerabilities.

## MATERIALS AND METHODS

### Data Source and Study Population

Our dataset comprises data for 5,857 patients diagnosed with PCa that accessed any of nine ECU Health hospitals between 2005 and 2025. Patient data included de-identified ECU Health System cancer registry records abstracted at ECU Health Medical Center, with analytic cases defined as patients diagnosed and/or treated at one of the nine hospitals. Some patients were analytic at duplicate hospitals if they received a portion of their primary diagnosis and/or therapy at more than one hospital: in total, the dataset contains ≤ 5.0% of registries with the same values in the fields of “Date of Diagnosis”, “Age at Diagnosis”, “Race”, and “Ethnicity”. This study was approved by the University and Medical Center Institutional Review Board at ECU Health.

For primary analysis we examined the AJCC summary stage of cancer at diagnosis and other known prognostic factors for PCa, including pre-treatment PSA levels, patients age distributions, Gleason score (GS), hospital location, diagnosis year, county of residence, self-declared ethnicity, and observed survival/death rates.

### Statistical Analysis

All statistical analyses were performed in *Python* (version 3.12.2). Comparisons between distributions were performed using Mann-Whitney (MW) non-parametric tests as implemented in the *SciPy* ^17^ package (version 1.17.1). Significance threshold was set to α = 0.05 modified using a Bonferroni correction to account for multiple hypothesis testing. Effect sizes were analyzed using the Common Language Effect Size (CLES) *f*. In this context, *f* equals the probability of a random element from one group having a higher value than a random element from the second group. To study the distribution of times from diagnostic to death we performed fitting of uncensored data and goodness-of-fit testing to exponential distributions using the Anderson-Darling test implemented in the *SciPy* package. Kernel density estimates for distributions were plotted using the *Seaborn* ^18^ package (version 0.13.2). Values are reported as mean ± standard error unless otherwise noted.

Survival ratios were calculated using the Kaplan-Meyer (KM) estimator and differences in survivability where quantified using a log-rank test as implemented in the *lifelines* ^19^ package (version 0.30.3).

## RESULTS

Among 5,857 cases from 2005 to 2025, 51.4% were self-identified White, 47.4% Black, and 0.8% other. Race was not self-identified in ≤ 0.5% of patients. The median age at diagnosis was 66 years. Sixty-three percent (63.3%) of patients were analytic from ECU Health Medical Center, while 36.7% of patients were from eight community partner hospitals. Analytic patients came from all 29 counties, ranging in numbers from N=1 (Columbus County) to 1395 patients (Pitt County), with 10 counties accounting for 74% of patients.

### Prognostic Factors

Analysis by stage revealed that the majority of patients in the dataset were stage 2 PCa, followed by stage 3, stage 4 and stage 1, (**Error! Reference source not found**.) with 20.2% of the patients not having a cancer stage assigned in the dataset. Temporal patterns occurred with staging. In 2005, 82.7% of the patients with documented stage were stage 2 cancers, and that proportion decreased to 43.7% by 2024. This large decrease was accompanied by an increase in the other stages: stage 1 new diagnoses increased from 1.8% in 2005 to 12.1% in 2024; stage 3 new diagnoses increased from 8.9% in 2005 to 28.2% in 2024, and stage 4 new diagnoses increased from 6.5% to 15.9% in the same period (Figure 1).

**Figure 1.**
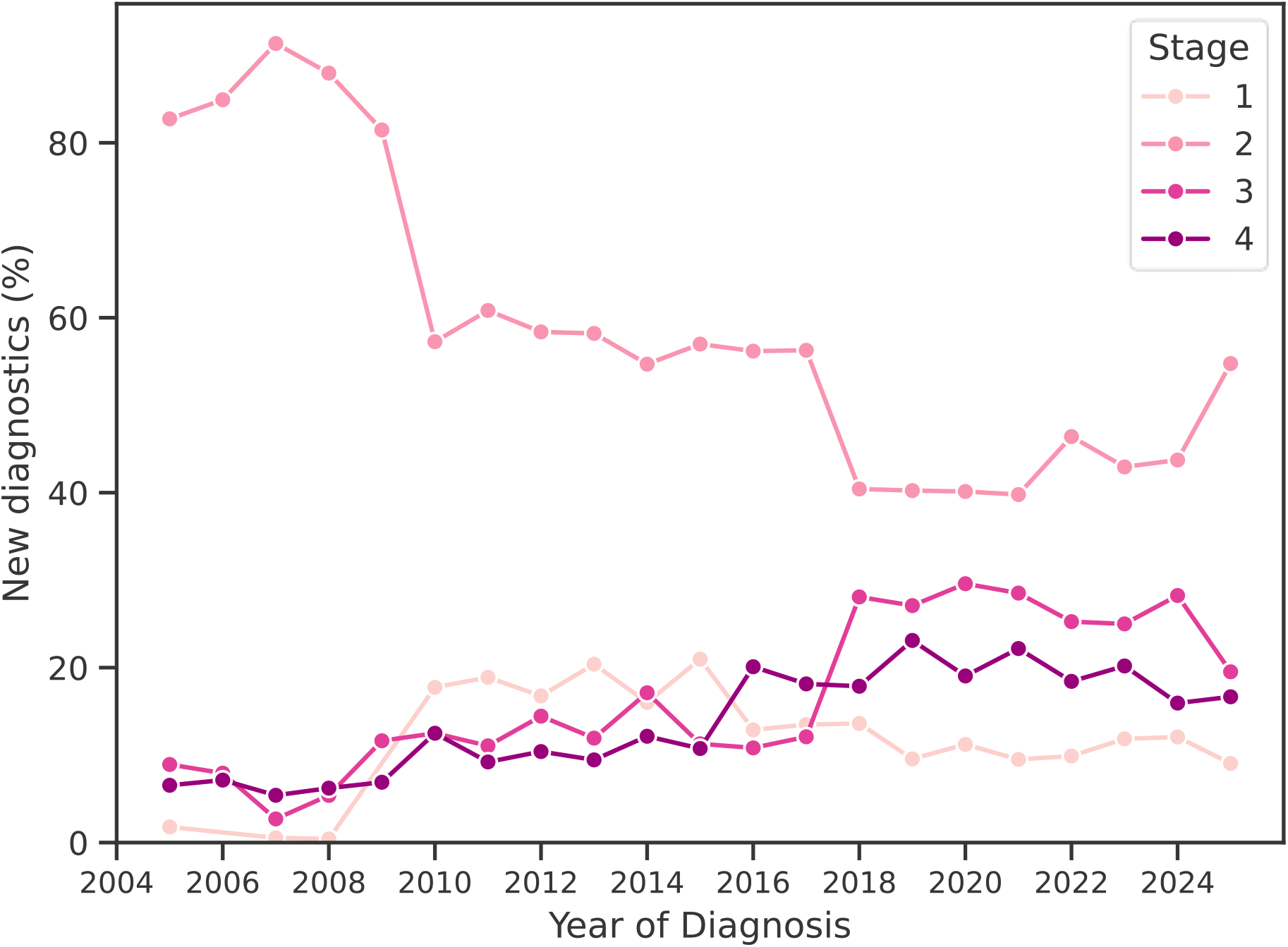
Proportion of new diagnoses of PCa by stage from 2005 to 2025.

PSA levels are also associated with PCa stage (Figure 2), with higher PSA levels corresponding to later stages (**Error! Reference source not found**.). Differences between PSA levels for the four stages were all significant (*P* < 0.001, *f* = 0.19 ± 0.04).

**Figure 2.**
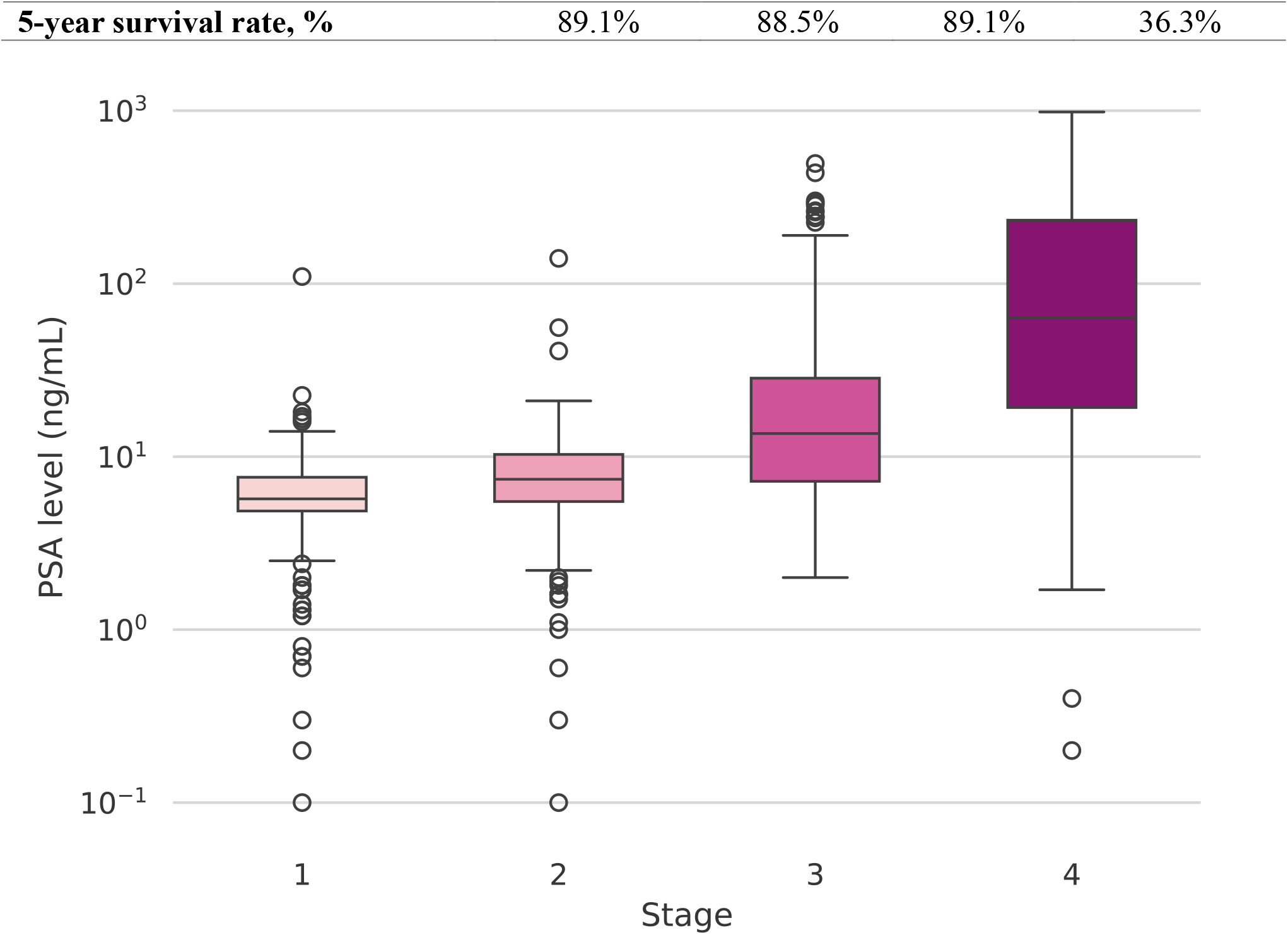
PSA levels of patients according to their cancer stage. Values for patients in the second and third quartile are enclosed by each box, while whiskers extend to include values in the first and fourth quartile. Points beyond 1.5 times the Interquartile Range (IQR) are considered outliers and are depicted in the figure by empty circles.

Patients presented with GS ranging from 3-10 over the period. The median Gleason score was GS 7 but varied temporally. Rare patients with GS < 5 occurred in later years (<0.3%). No significant differences in GS occurred between Black and White patients.

### Age Distribution of Patients

Age distributions were also distinct for each of the PCa stages: stages 1, 2 and 3 have similar means and standard deviations (Table 1). Stage 4, however, displayed an elevated mean and standard deviation (Figure 3). Pairwise MW tests with Bonferroni correction uncovered no significant differences among stages 1, 2 and 3 (1 vs. 2: *P* = 0.787, *f* = 0.50; 1 vs. 3: *P* = 0.440, *f* = 0.49; 2 vs. 3: *P* = 0.465, *f* = 0.49). Stage 4 differed significantly from all three remaining stages (1 vs. 4: *P* < 0.001, *f* = 0.37; 2 vs. 4: *P* < 0.001, *f* = 0.38; 3 vs. 4: *P* < 0.001, *f* = 0.38). It is worth noting that 15.7% of all patients presented at diagnosis above the age of 75 (stage1: 12.7%, stage 2: 12.1%, stage 3: 12.3%, stage 4: 31.0% of the patients). The proportion of patients being diagnosed at the age of 70 is even larger: 30.9% of the patients were diagnosed at the age of 70 or above (stage 1: 28.4%, stage 2: 28.1%, stage 3: 26.1%, stage 4: 47.2% of the patients, see Supplementary Figure SF1).

**Table 1.** Characteristics of the patients by stage.

|  | **Stage 1** | **Stage 2** | **Stage 3** | **Stage 4** |
|---|---|---|---|---|
| **Patients, n (%)** | 517 (8.8) | 2,632 (44.9) | 851 (14.5) | 675 (11.5) |
| **Black** | 251 (9.0) | 1217 (43.8) | 420 (15.1) | 374 (13.5) |
| **White** | 280 (9.3) | 1502 (49.9) | 461 (15.3) | 322 (10.7) |
| **Age at diagnosis, mean ± std. err.** | 65.4 ± 0.4 | 65.6 ± 0.2 | 65.9 ± 0.3 | 70.1 ± 0.4 |
| **Black** | 63.6 ± 0.5 | 64.4 ± 0.2 | 64.1 ± 0.4 | 68.3 ± 0.5 |
| **White** | 67.2 ± 0.5 | 66.7 ± 0.2 | 67.5 ± 0.4 | 72.2 ± 0.6 |
| **PSA levels, mean ± std. err.** | 6.6 ± 0.5 | 8.5 ± 0.2 | 28.0 ± 1.9 | 178.4 ± 13.0 |
| **Gleason score, median (IQR)** | 6 (0) | 7 (0) | 7 (1) | 8 (2) |
| **Median survival time, years** | 17 | 17 | 19 | 3 |

**Figure 3.**
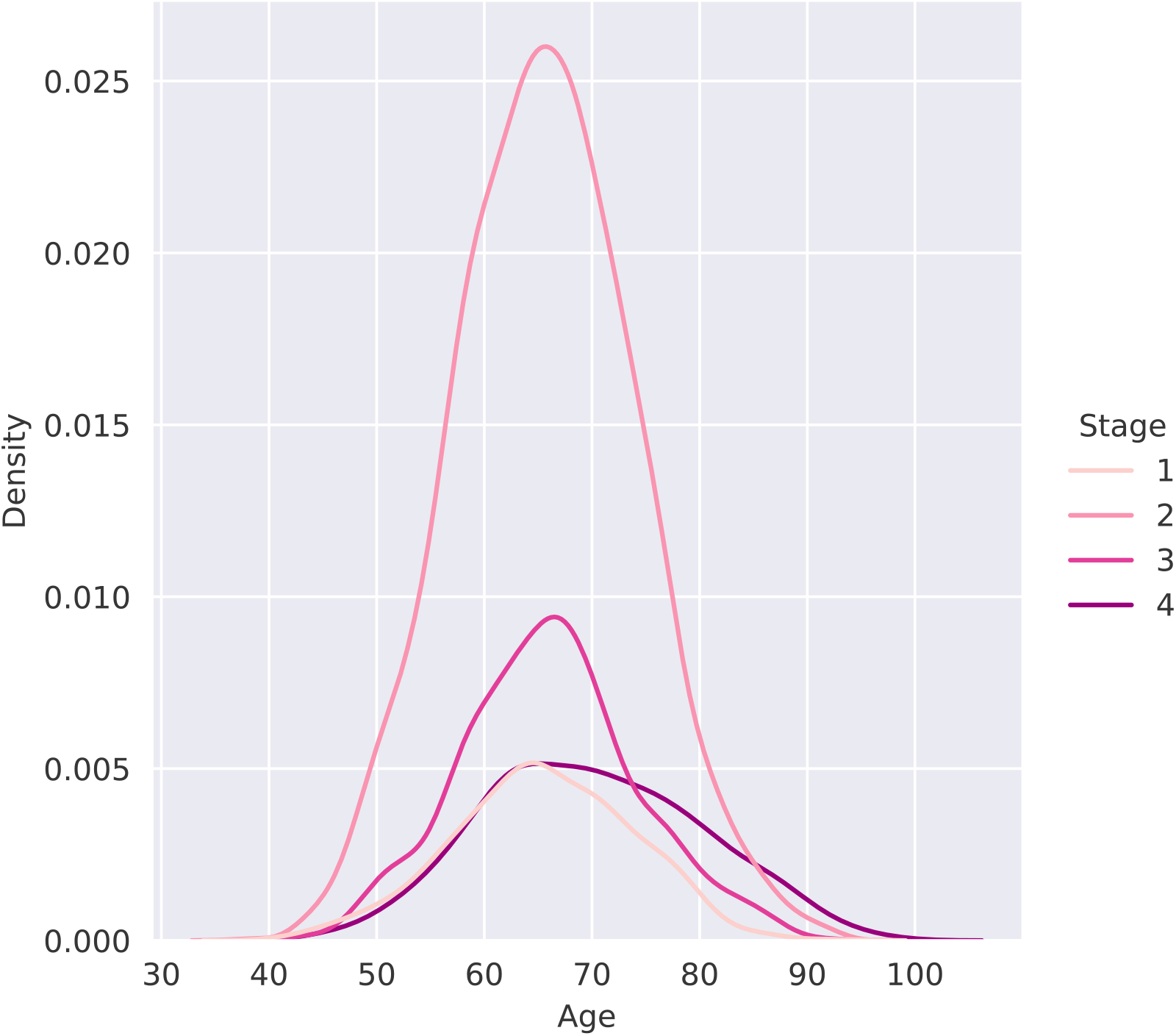
Kernel density estimates of the age distribution of patients according to their diagnosed stage.

Age distributions slightly shifted toward older ages from 2005 (65.3 ± 0.6 y) to 2025 (69.0 ± 0.5 y). Significant differences (Supplementary Figure SF2) only begin to appear between age distributions that are at least 7 years apart (largest significance: 2010 vs. 2024, *P* < 0.001, *f* = 0.36; smallest significance: 2014 vs. 2021, *P* < 0.001, *f* = 0.41), with the proportion of comparisons yielding significant differences increasing as the difference between years increases (7 years difference: 21.4% of comparisons yield significant differences; 14 years difference: 71.4% of all comparisons display significant differences; 20 years difference: 100.0% of comparisons display significant differences). Effect sizes are small or negligible except for five cases (2006 vs. 2025, *P* < 0.001, *f* = 0.33; 2008 vs. 2025: *P* < 0.001, *f* = 0.34; 2010 vs. 2025, *P* < 0.001, *f* = 0.34; 2012 vs. 2025: *P* < 0.001, *f* = 0.34; and 2006 vs. 2024, *P* < 0.001, *f* = 0.35) where they are medium ^20^.

Black patients present consistently at a significantly younger age than White patients (Table 1) for all stages of PCa (stage 1: *P* = 0.001, *f* = 0.37; stage 2: *P* < 0.001, *f* = 0.42; stage 3: *P* < 0.001, *f* = 0.38; stage 4: *P* < 0.001, *f* = 0.38), yet they present a higher number and proportion of stage 4 PCa in the latter years (Figure 4).

**Figure 4.**
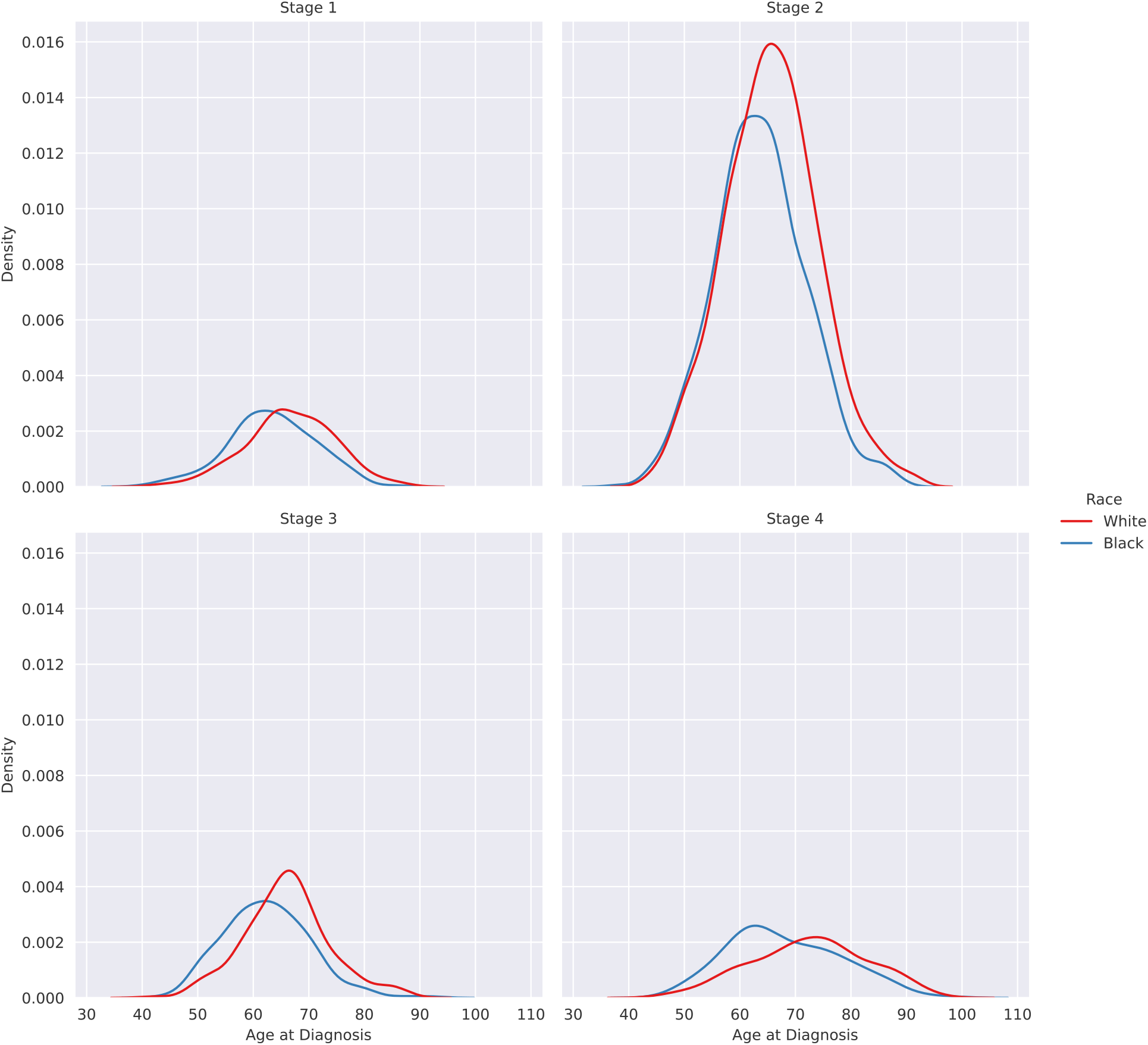
Kernel density estimates of age distributions of patients according to their race and stage diagnosis.

### Survival rates

Probability of survival depends on stage (Figure 5). Median survival times for stage 1, 2 and 3 patients are 5 times higher than for stage 4 (**Error! Reference source not found**.). The survival function for stage 4 was significantly different than for the rest of stages (*P* < 0.001). The 5-year survival rate further evidences the singularity of stage 4 PCa: survival rates for stage 1,2 and 3 patients are almost three times higher than for stage 4 patients (**Error! Reference source not found**.). Observed survival rates are similar between Black and White patients (Supplementary Figure SF4).

**Figure 5.**
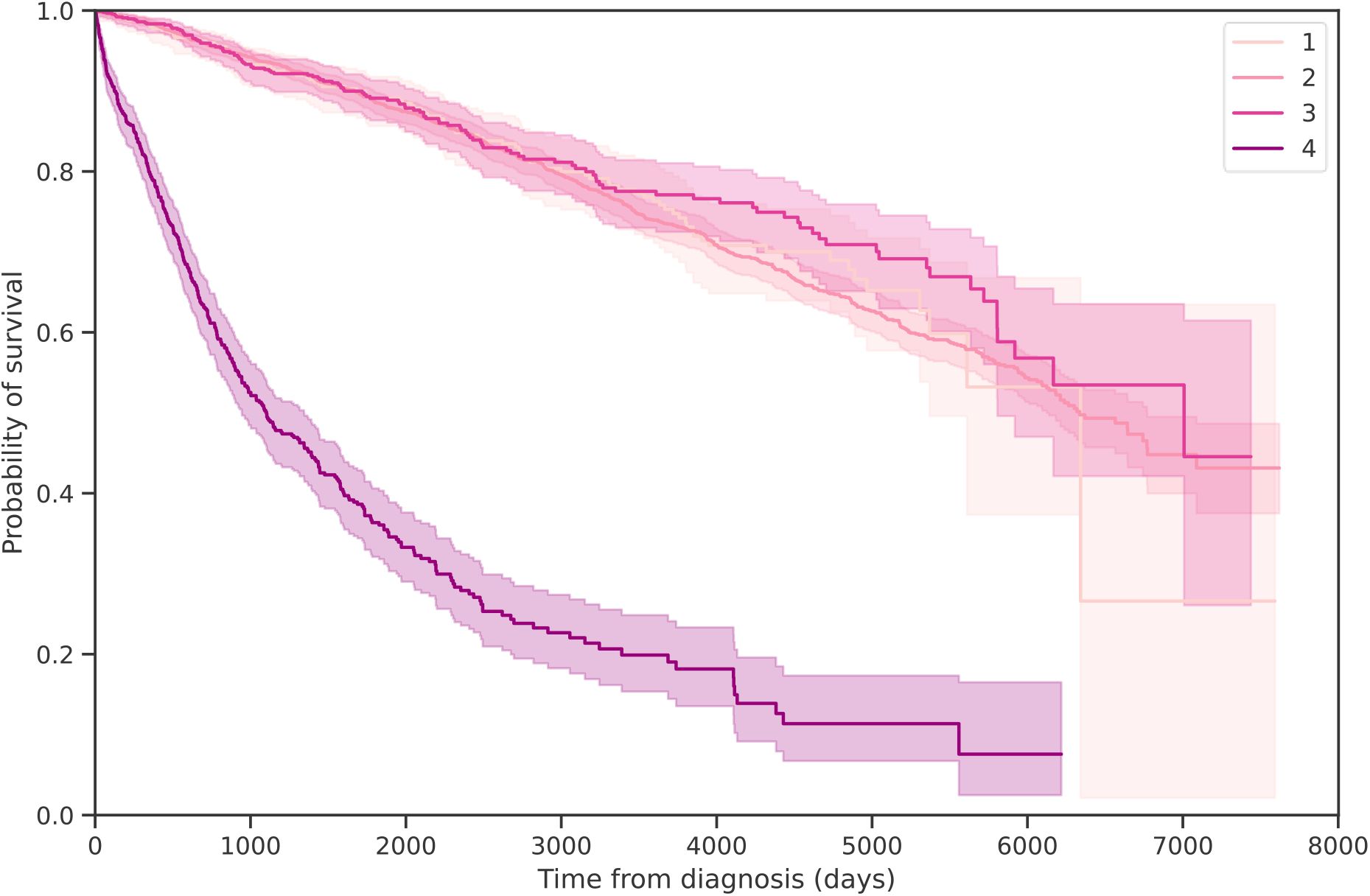
Kaplan-Meyer plot of the survival function by stage. Shaded area represents 95% confidence interval.

Even though the distribution of time from diagnosis to death for those patients that died for stages 1, 2 and 3 do not resemble any known distribution, the time from diagnosis to death for patients with stage 4 PCa follows an exponential distribution (*β* = 30.9, *A*^*2*^ = 1.333, *P* ≤ 0.05; see Supplementary Material for a complete discussion).

### Hospital Location and Size

By hospital catchment area, diagnoses are not equitably distributed. Smaller hospitals see a disproportionate number of late-stage PCa when compared to larger hospitals (Figure 6): while in the central hospital the proportion of stage 4 is 12.5%, the proportion of stage 4 increases to 33.3% ± 9.1% in the smaller community hospitals. Patients accessing community hospitals are also more likely to experience an increase in their GS following primary surgery (pathologic grade higher than clinical grade): 20.8% of patients accessing the community hospitals had their GS increased after surgery, compared to 14.8% of patients accessing the central hospital in the network (see Supplementary Figure SF5). Survival odds are also different when comparing the central hospital with the community hospitals (Supplementary Figure SF7): while the 5-year survival odds for the central hospital are 83.7%, for community hospitals the odds decrease to 77.6% (*P* < 0.001).

**Figure 6.**
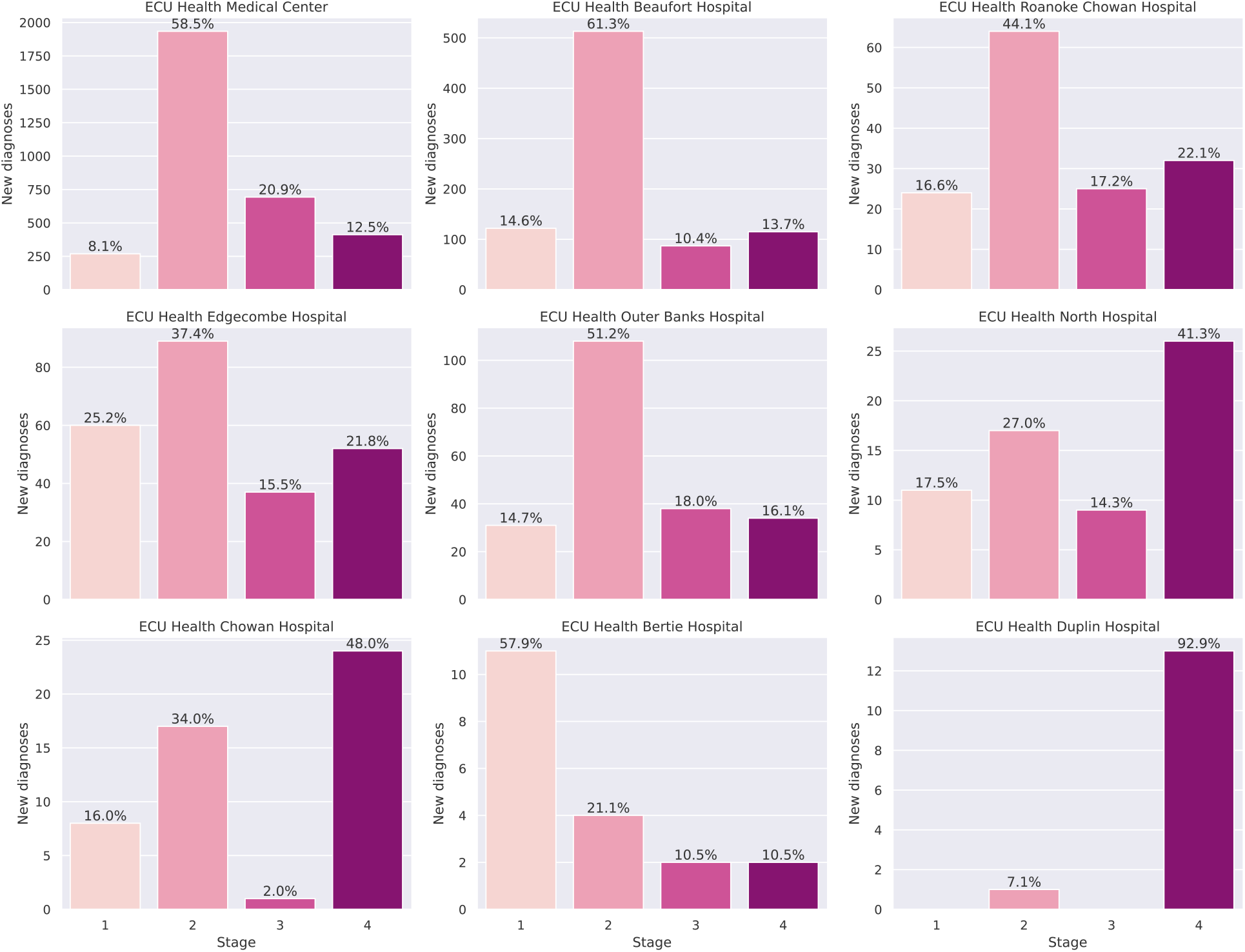
Volume and proportion of new diagnoses by hospital and stage. Each panel corresponds to a different hospital, and each bar represents the new diagnoses of each stage. Percentages for each stage are displayed on top of the bars. Hospitals are presented in order of total volume of new diagnoses (left to right and top to bottom).

## DISCUSSION

Our analyses describe patterns in PCa in the 2005-2025 period within a 9-hospital network in rural ENC. The changes in the incidence of by stages over a twenty-year period are a consequence of several occurrences which culminate in two inflection points (Figure 1). First, in 2010, stage 2 cases drop precipitously as stages 1, 3 and 4 increase. This change reflects the inclusion of better prognosticators in staging (Gleason score consensus, inclusion in stage) and improved imaging modalities (MRI of prostate, PSMA PET scan) which now allow better prognostication and risk stratification into stages ^6^. At the same time, new guidelines from the US Preventive Services Task Force (USPSTF) in 2012 recommended against PSA-based screening for PCa ^21^. Although the USPSTF was criticized for underestimating the benefits of PCa screening while overestimating the risks ^22^, its decision effectively decreased the diagnoses of PCa especially in rural areas where patients have limited access to specialists ^23^. In 2018, the USPSTF changed its guidelines and endorsed individual decision-making ^24^. This resulted in an overall increase in the number of cases ^25^, which led to the second inflection point in our system. Starting in 2018 our data evidences an increase particularly in stages 3 and 4 cases, accompanied by a decrease in stages 1 and 2 cases, confirming the effectiveness of an active surveillance system despite the fewer options to have detailed discussions on screening.

It is possible that changes observed over time in the dataset are a result of the aging patient population, and, in fact, our analyses reveal that the age distribution of patients with PCa has shifted towards older ages, which is consistent with current demographic trends ^26^. However, the small effect sizes found in our analyses when comparing age distributions and the fact that significant differences in age distributions of patients can only be observed in 7-year span periods or above, indicate that differences in age distributions alone cannot explain the differences observed in the dataset.

Nevertheless, we found a significant difference with medium effect size between the age distribution of patients with stage 4 PCa and the rest of the patients. These findings suggest that stage 4 patients constitute a distinct population with substantially older patients, which might indicate a failure to detect the cancer at an earlier stage and age. Our results are consistent with national trends of increasing distant disease at diagnosis for men over age 75 following current screening recommendations ^27^. Since the ability to detect cancer at an earlier stage is conditioned by a patient’s knowledge about screening, access to healthcare providers for screening, and transportation to appointments ^28^, this failure to detect PCa at an earlier stage might be evidencing limitations in access to early detection. Furthermore, current recommendations for PCa screening (stop screening at 75 y) leave 12% of the population with non-stage 4 PCa out of the screening process (27% if the recommendation followed is to stop screening at the age of 70 y), which leads to patients with undetected PCa presenting with a late-stage PCa at a later age.

The connection between rural areas and limited access to testing is supported by our analysis of hospital sizes. The results of those analyses evidence that community hospitals in predominantly rural areas see a disproportionate amount of stage 4 PCa cases when compared to the central hospital, which might indicate a lack of adequate access to primary care or specialists in the area. In rural areas where the hospital serving that community is small and does not have a specialist, patients may not request or obtain regular testing in the primary care setting, and only present to their nearest hospital when the pain or discomfort levels from an advanced cancer are severe.

The existence of disparities between rural and urban hospitals is also confirmed by the observed changes in GS after surgery: patients from rural areas presenting at community hospitals are more likely to see their GS increased after primary surgery. A plausible explanation for this difference is that there is a bias towards lower clinical GS in smaller community hospitals, probably due to the lack of adequate testing equipment or specialists. This bias is then corrected after primary surgery, which explains the difference in increase in GS between patients accessing community hospitals and patients accessing the central hospital. One of the consequences of these disparities between central hospital and community hospitals can be found in the survival ratio of patients, which is significantly lower for patients accessing the community hospitals.

The importance of early detection can be seen in Figure 5, where the odds of survival in the first 5 years after diagnosis are 6 times higher for non-stage 4 PCa than for stage 4 PCa. For stage 4 PCa, the treatment options are very limited and frequently involve palliative care only, which leads to worst outcomes for patients ^29^. Moreover, the time span from diagnosis to death for those patients diagnosed with stage 4 PCa that died after diagnosis follows an exponential distribution. In this context, our results indicate that deaths after stage 4 PCa diagnosis occur continuously and at a fixed common average rate for all patients. Furthermore, since exponential distributions are memoryless, the probability of dying only depends on the timespan under consideration regardless of the time since diagnosis.

Demographic distribution of patients across geography can also be a contributing factor to the differences observed: smaller community hospitals in rural areas tend to have a predominantly Black patient population. It is worth noting, though, that while detection of PCa happens at an earlier age for Black patients than for White patients for all stages of PCa, a higher proportion of Black patients are diagnosed with late-stage PCa. It is unclear from the data what is driving this inequity between Black and White patients, and a deeper analysis using other social drivers of health should be carried out in order to extract accurate conclusions. No differences in survival rates between Black and White patients are observed.

Limitations of this study include changes in staging that are not fully reflected in the cancer registry. Even though AJCC staging has recommended stage 4A and 4B since the 6^th^ edition in 2004 ^30^, many rural providers are still using generic stage 4, so subset analysis is problematic. Furthermore, stage information was lacking or unknown for 966 patients, representing nearly 20% of the population. However, survival analysis of these patients shows 5-year results consistent with early-stage patients, suggesting they were not stage 4.

## CONCLUSIONS

Our results evidence the complexity of healthcare in predominantly rural areas and suggest that rural healthcare access is a potential driver of disparities. Smaller hospitals serving rural communities saw a substantially higher proportion of late-stage PCa diagnoses and lower survival rates than larger hospitals, consistent with limited local access to specialists and routine screening. These findings suggest that health systems serving rural communities should consider targeted interventions to reduce the burden of late-stage diagnosis in underserved populations.

## Data Availability

Non-identifiable data is available from the authors upon reasonable request and with permission of ECU Health.

