## Supplemental material for "Challenges and Health Disparities in Prostate Cancer: Healthcare Access as a Driver of Inequities in a Rural Hospital Network"

Supplementary material

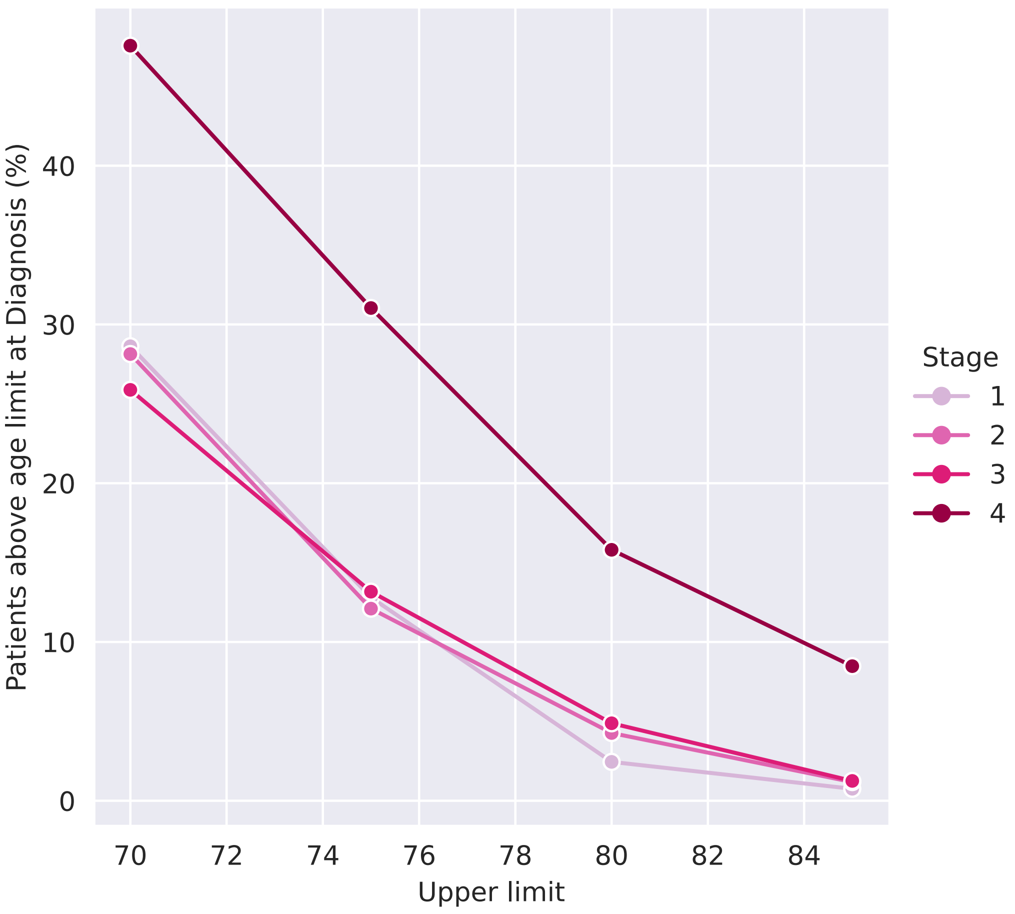


**Supplementary Figure SF1**. Proportion of patients above a certain age at the time of diagnosis. Each line corresponds to a different diagnosed stage of PCa.

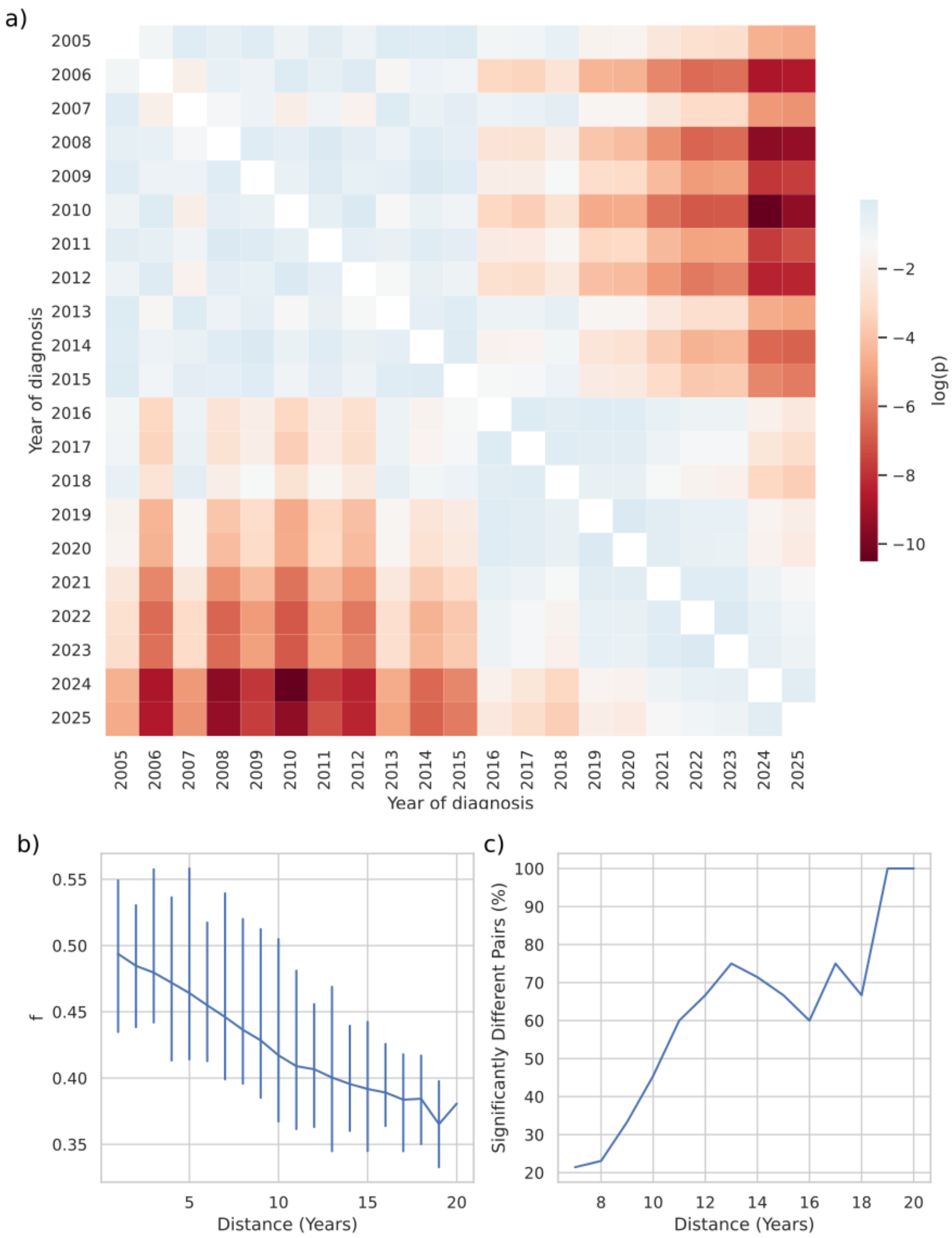


**Supplementary Figure SF2.** Comparison of age distributions of different years. **a)** Results of pairwise MW tests between age distributions. Each cell on the heatmap is colored according to the p-value (in logarithmic scale) of the MW test of the corresponding years under comparison. **b)** Effect sizes calculated as CLES according to the distance (timespan) between the years under comparison. The line depicts the mean, while vertical bars include the values for all the comparisons in that distance. **c)** Proportion of comparison yielding significant differences according to the distance between the years under comparison.

### Distribution of timespan between diagnosis and death

We fitted the distribution of timespans from diagnosis to death using a normal distribution (using the D'Agostino-Pearson test), a lognormal distribution (using the D'Agostino-Pearson test), and an exponential distribution (using the Anderson-Darling test). For numerical stability, we substituted all values < 0 by 1. In the D'Agostino-Pearson test, a statistic of 0 is associated with a perfect fit to a normal (or lognormal) distribution, with the p-value indicating the probability of obtaining that statistic from a normal (or lognormal) distribution. In the Anderson-Darling test, values for the statistic are compared to pre-established charts that depend on the distribution to fit (exponential), with *P*-values indicating the confidence with which the null hypothesis (that the data is pulled from an exponential distribution) can be rejected.

**Supplementary Table ST1**. Goodness-of-fit of different distributions to the distribution of timespans from diagnosis to death.

| | Normal fit | | Lognormal fit | | Exponential fit | |
|---|---|---|---|---|---|---|
| | Statistic | *P*-value | Statistic | *P*-value | Statistic | *P*-value |
| Stage 1 | 4.3138 | 0.1157 | 31.2933 | < 0.0001 | 3.6490 | > 0.85 |
| Stage 2 | 67.0724 | < 0.0001 | 241.5072 | < 0.0001 | 35.6938 | > 0.85 |
| Stage 3 | 12.7056 | 0.0017 | 25.3858 | < 0.0001 | 2.6129 | > 0.85 |
| Stage 4 | 123.0484 | < 0.0001 | 30.0385 | < 0.0001 | 1.3329 | ≤ 0.05 |

We compared the goodness-of-fit of a powerlaw distribution to the fitted lognormal distribution and to the fitted exponential distribution. Using the methodology described in [1], positive values of the statistic R indicate a better fit using a powerlaw than using the distribution under comparison, while p-values indicate the significance value for that direction.

**Supplementary Table ST2**. Comparison of powerlaw fitting to other distributions.

| | Powerlaw vs lognormal | | Powerlaw vs. exponential | |
|---|---|---|---|---|
| | Statistic (R) | *P* | Statistic (R) | *P* |
| Stage 1 | -2.8849 | 0.0039 | -4.9508 | < 0.0001 |
| Stage 2 | -8.0087 | < 0.0001 | -14.2577 | < 0.0001 |
| Stage 3 | -1.8010 | 0.0717 | -2.4414 | 0.0146 |
| Stage 4 | -3.7870 | 0.0002 | -3.7826 | 0.0002 |

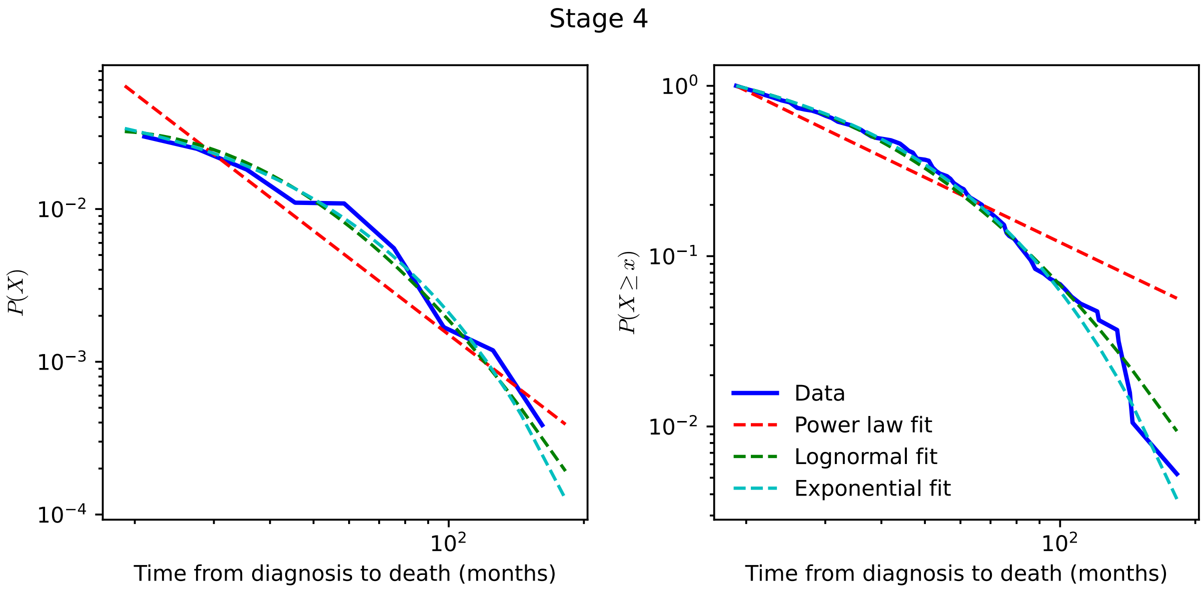


**Supplementary Figure SF3**. Comparison of three different models with the data in the dataset (solid blue line): a power law (dashed red line), a lognormal (dashed green line), and an exponential (dashed cyan line). Left: probability density function. Right: Complementary cumulative density function.

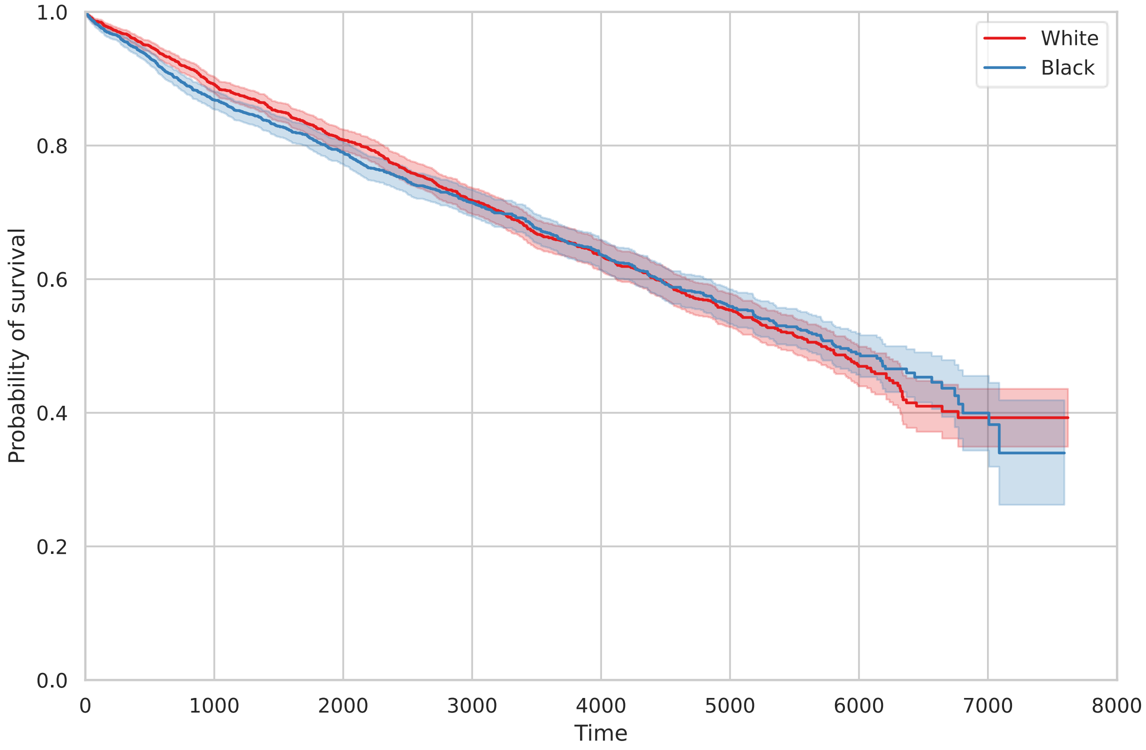


**Supplementary Figure SF4**. Kaplan-Meyer plots of the survival function. Each line corresponds to a different ethnic group. The log-rank test displays no difference in survival rates between Black and White patients ($P = 0.6239$).

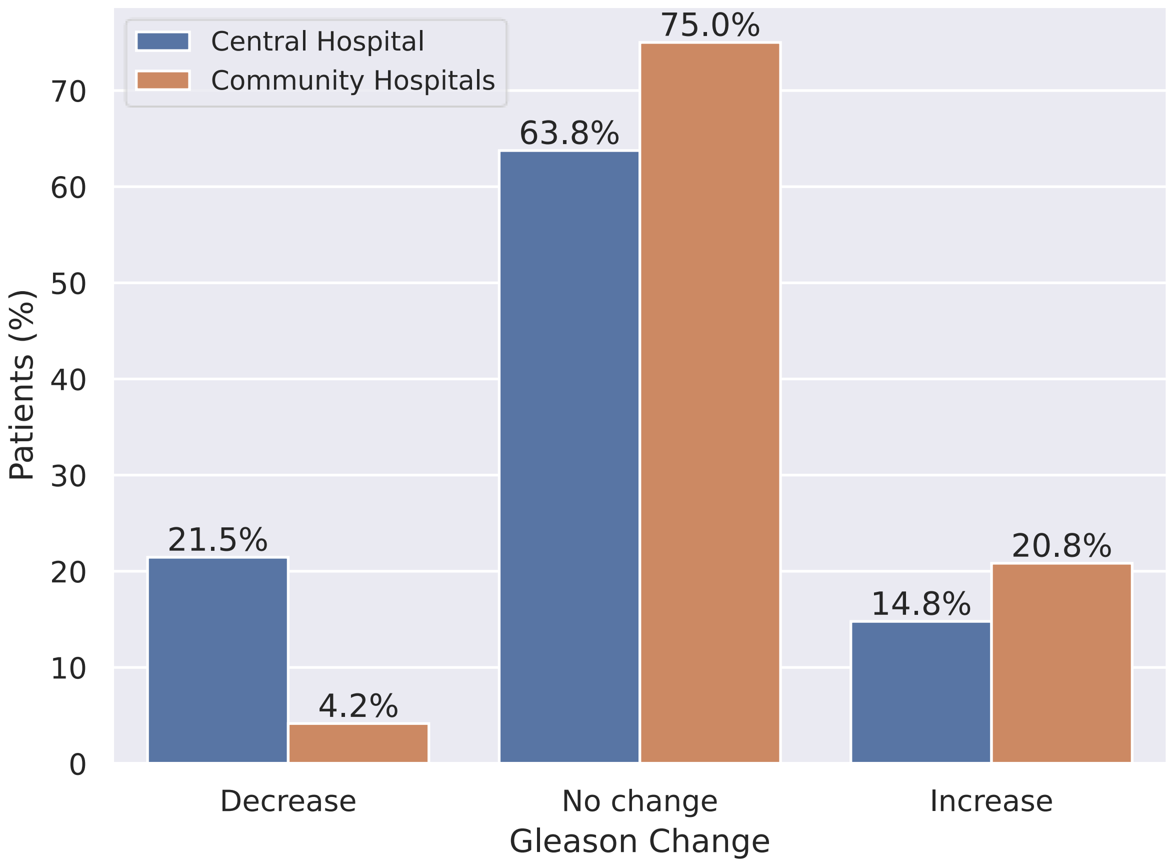


**Supplementary Figure SF5**. Proportion of patients that have their Gleason score increased, decreased or unchanged after surgery, according to the hospital of presentation: blue bars represent patients in the central hospital, while brown bars represent patients in one of the eight community hospitals. Numbers above bars represent percentages of patients in the hospital group.

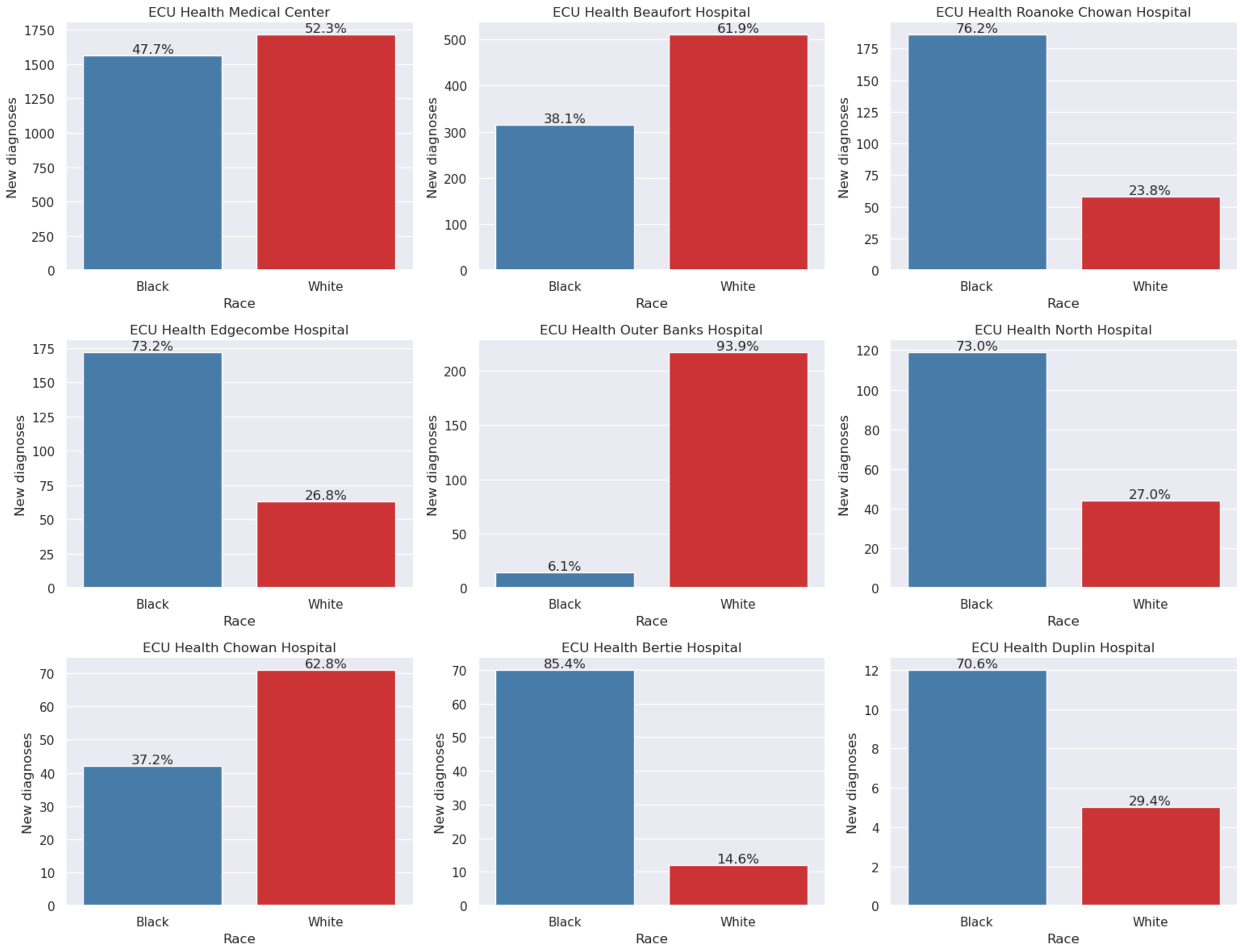


**Supplementary Figure SF6.** Number of patients by race diagnosed with PCa at each hospital of the network. Numbers above bars represent percentages of each race.

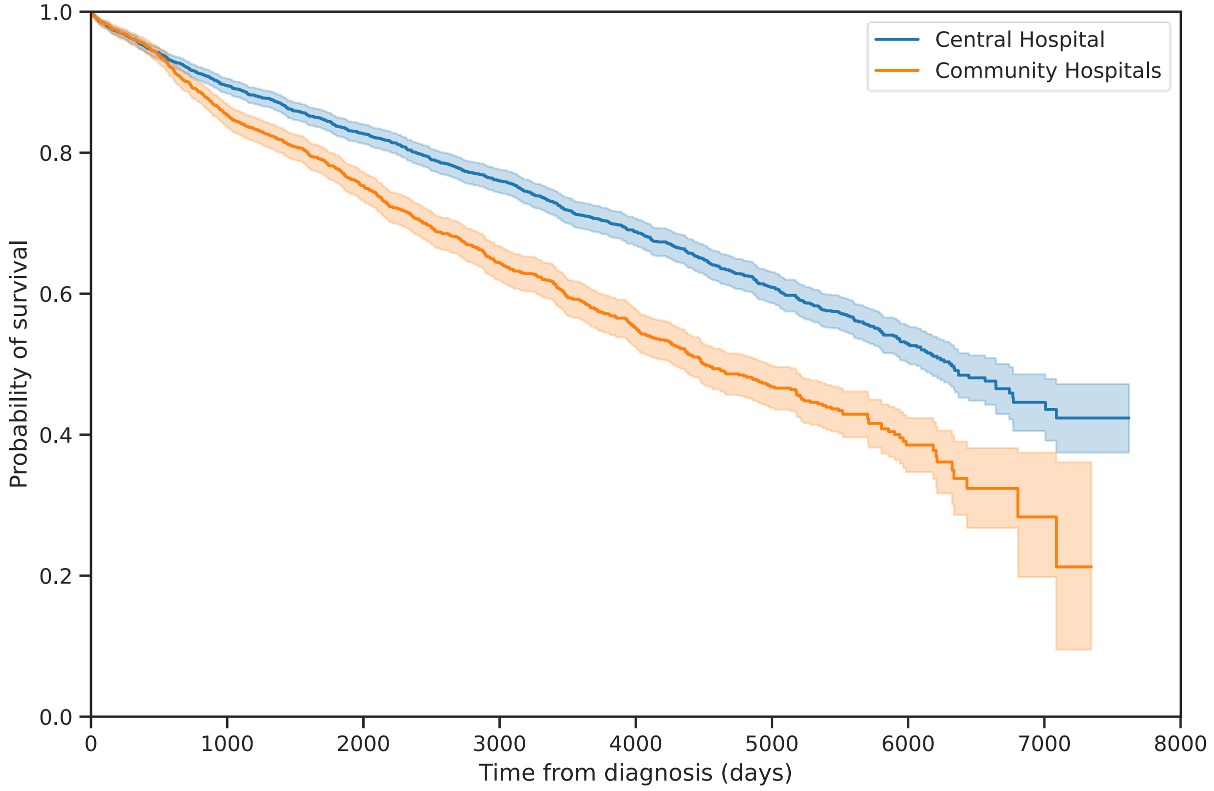


**Supplementary Figure SF7**. Kaplan-Meyer plot of the survival probability of PCa patients according to the hospital they access. Shaded area represents 95% confidence interval.
